# All You Need Is Context: Clinician Evaluations of various iterations of a Large Language Model-Based First Aid Decision Support Tool in Ghana

**DOI:** 10.1101/2024.04.03.24305276

**Authors:** Paulina Boadiwaa Mensah, Nana Serwaa Quao, Sesinam Dagadu, James Kwabena Mensah, Jude Domfeh Darkwah, Project Genie Clinician Evaluation Group [1]

**Affiliations:** SnooCODE RED, SnooCODE, Accra, Ghana; Accident and Emergency Centre, Korle Bu Teaching Hospital, SnooCODE RED, Accra, Ghana; Volta Regional Government Hospital, Hohoe, Ghana; University Hospital, Kwame, Nkrumah, University of Science and Technology, Kumasi, Ghana; Ghana

**Keywords:** SnooCODE, Clinical Decision Support, Large Language Models, First Aid, Emergency Medical Services, Medical Emergencies, Clinical Context, Clinician Evaluation, Resource-Constrained Settings, Gemini 1.5 Pro, GPT 4, Claude Sonnet

## Abstract

As advancements in research and development expand the capabilities of Large Language Models (LLMs), there is a growing focus on their applications within the healthcare sector, driven by the large volume of data generated in healthcare. There are a few medicine-oriented evaluation datasets and benchmarks for assessing the performance of various LLMs in clinical scenarios; however, there is a paucity of information on the real-world usefulness of LLMs in context-specific scenarios in resource-constrained settings. In this work, 5 iterations of a decision support tool for medical emergencies using 5 distinct generalized LLMs were constructed, alongside a combination of Prompt Engineering and Retrieval Augmented Generation techniques. 50 responses were generated from the LLMs. Quantitative and qualitative evaluations of the LLM responses were provided by 13 physicians (general practitioners) with an average of 3 years of practice experience managing medical emergencies in resource-constrained settings in Ghana. Machine evaluations of the LLM responses were also computed and compared with the expert evaluations.

## I. Introduction

***“Provide a cool mist humidifier or take the infant into a steamy bathroom to help loosen mucus.”* –** this was First Aid Step no.3 provided by Claude 3 Sonnet for managing possible Bronchiolitis or Asthma Exacerbations – two conditions that cause breathing problems. While this may be valuable advice, it might not be applicable to a child living on a rural cattle farm in Akobo, South Sudan. When this particular location is added to the prompt, the response makes no mention of mist humidifiers and steamy bathrooms. Rather the first step provided by the model is to ***“Move the infant to an area with fresh air and away from any dust/irritants.”*** This shows the importance of considering the background contexts of prompts in evaluating the performance of Large Language Models (LLMs). Amongst the popular biomedical Natural Language Processing (NLP) datasets for evaluating LLMs, none of them have been specifically prepared for resource-constrained settings as found in Low-and Low-Middle-Income countries (LMICs) [2]. Thus, though a few models achieve high scores when evaluated on these datasets, their translational value in everyday clinical scenarios in LMICs cannot be readily ascertained. In this work we aim to add to the limited knowledge base on LLM applications for clinical scenarios in LMICs. Specifically, we aim to evaluate the appropriateness of some selected generalized LLMs for use in clinical decision support tools in LMICs and to provide a reference for future, more expansive research. After conducting several experiments, we found that when generalized LLMs are given prompts that aim to generate first aid advice for medical emergencies, their outputs differ significantly when additional context-specific location is provided [3]. Thus, we provided context-specific prompts and asked clinicians with substantial familiarity with those contexts and clinical scenarios to evaluate the outputs. This work is part of a research and development process to eventually deploy LLM-based Clinical Decision Support tools for managing medical emergencies in resource-constrained settings.

## II. Related Work

Prior studies have shown that though there are vital concerns to be addressed, the general consensus is that LLMs hold immense potential in improving healthcare delivery when they are incorporated in various capacities such as: in automation of administrative tasks, clinical decision support tools, virtual health assistants, screening tools, health trackers, clinical language translation tools, medical research and health education tools [4][5][6][7]. These use cases can augment the limited financial, logistical, and human resources available in LMICs [8]. Initial studies on clinician perception on the usefulness of a combination of OpenAI’s “gpt-3.5-turbo / “gpt-4” and Retrieval Augmented Generation (RAG), as a health education tool in India, an LMIC, revealed that though clinicians believed the tool held potential, they were generally not satisfied with its performance [9]. In that study, the authors identified the need to enhance the contextual and cultural relevance of the models’ responses. Another comparative study of a clinician evaluation of Almanac, an LLM framework based on OpenAI’s “text-davinci-003” combined with RAG, versus ChatGPT reveals that though clinicians rated Almanac’s answers as safer and more factual, they still preferred ChatGPT’s answers [10]. However, this study does not reveal whether the clinicians shared their perspective on the usefulness of any of the models for everyday clinical scenarios, neither does it capture the perspectives of clinicians who practice in LMICs.

## III. Methodology

### A. LLM Selection

We selected Open AI’s GPT-4 Turbo Preview, both via the Assistant Application Programming Interface (API) and the Chat Completions API. We evaluated these separately as the temperature of the model was almost impossible to be tweaked when using the OpenAI Assistant, at the time of the study. In addition, we selected Gemini 1.5 Pro and Claude Sonnet. These models were selected based on performance on popular benchmark evaluation tests [2], their ranking on the LMSYS Chatbot Arena Leaderboard as of 13th April 2024. The selected models were top performers on the leaderboard (in Ranks 1 to 5) [11]. Furthermore, we factored in the availability of API, and ease of access in selecting models. We did not select similarly ranked open-source medical LLMs because of the computational resources required to run/access them, for example, advanced GPUs. We then tested a combination of prompt-engineering and Retrieval Augmented Generation (RAG) techniques to produce outputs/responses from the various LLMs as follows:

- GPT 4-Turbo Preview via Open AI Assistant API + Prompt Engineering = ***Response A***
- Gemini 1.5 Pro + Prompt Engineering = ***Response B***
- Claude Sonnet + Prompt Engineering = ***Response C***
- GPT4-Turbo Preview via Open AI Chat Completions API + Prompt Engineering + RAG = ***Response D***
- Claude Sonnet + Prompt Engineering + RAG = ***Response E***

### B. Parameter Tuning

The temperature was set at 0 for generating Responses C to E. This was to get deterministic responses as often as possible due to the critical nature of the proposed use case. For Response A, the default temperature used in Open AI Assistant was maintained as it was difficult to ascertain and tweak. For Response B, the default temperate of 2 set in the Google AI Studio was maintained as it was also difficult to tweak. An output length of 4000 was set in Google AI Studio for assessing Gemini 1.5 Pro to provide an ample window for the extent of generated responses. Similarly, the max tokens parameter was set at 4000 for assessing Claude Sonnet to provide an ample window for the extent of generated responses.

### C. Prompt Engineering

We employed in-context learning using one-shot inference. The prompt consisted of three parts, the system message/prompt/instructions, an example conversation and the input message. Here is an example of the input message for one of the prompts:

“

Location: rural area, Bongo, Ghana. There is a chemist 300m away and a district hospital 1km away. Patient’s age as: 5 months, sex as: male. Description of medical emergency: fall from stool, vomiting. 1. PATIENT CAN TALK NORMALLY 2. ATIENT CAN BREATHE NORMALLY 3. PATIENT HAS A NORMAL PULSE 4. PATIENT IS NOT VISIBLY BLEEDING 5. PATIENT IS AWAKE AND ALERT 6. PATIENT DOES NOT HAVE A VISIBLE TRAUMATIC INJURY, ANIMAL BITE OR RASH 7. PATIENT HAS NO KNOWN ALLERGIES 8. THE PATIENT HAS TAKEN PARACETAMOL 9. PATIENT

HAS NO KNOWN PAST MEDICAL HISTORY 10.THE TIME OF LAST MEAL WAS 30 MINUTES AGO

“

### D. Retrieval Augmented Generation

Retrieval Augmented Generation (RAG) has been touted as a highly promising approach to improving factuality, reasoning, and interpretability of LLM outputs [12][13]. We provided a free manual for first aid instruction geared towards settings in sub-Saharan Africa [14]. We employed two chunking approaches for implementing RAG. In the first, the text from the module was divided into chunks using the “CharacterTextSplitter” tool from Langchain. In the second, the text was divided into chunks using semantic splits via the “all-mpnet-base-v2” transformer model from the HuggingFace model hub. Vector embeddings were generated from the resulting chunks via the “text-embedding-3-large” embedding model from Open AI. This embedding model works well with both GPT4Turbo and Claude Sonnet for retrievals but does not work as well with Gemini 1.0 Pro, thus RAG was not tested with the Gemini model. The vector embeddings were stored in the open-source vector database, ChromaDB.

### E. Clinician Selection

Clinician evaluators were selected via a local clinician network, from diverse practice locations within Ghana and based on their familiarity with the locations, contexts, and clinical scenarios. Clinicians selected were verified to be in good standing with the Ghana Medical and Dental Council and had valid licenses to practice. Clinicians were asked to input their completed number of years of practice, and to round up surplus months to one year, for 10+ months and to round down to 0 years if less than 10 months. On average, clinician evaluators had 3 completed years of experience, as the first point-of-call in the hospital in managing medical emergencies in Ghana. It is expected that they possess sufficient knowledge and skills to deliver, at a minimum, first aid in the selected medical scenarios.

### F. Selection of Medical Scenarios

***“Love how it span over all major disciplines”*** – A clinician evaluator.

Ten simulated clinical scenarios were provided in the format shown in Section C above. The scenarios featured a wide range of demographics with the youngest simulated patient being 5 months old, and the oldest being 85 years old. The clinical scenarios cut across all major clinical specialties and were selected to reflect common emergency scenarios in Ghanaian health facilities. The simulated clinical scenarios are described in detail in the Google forms in this referenced folder [15]. There was an equal distribution of male and female patients in the scenarios represented.

### G. Response Evaluation and Ranking

There were 5 LLM responses to each of the 10 clinical scenarios making a total of 50 responses. These 50 responses were in two groups, in the first group, RAG was done with Approach 1 as described in Section D above. The first group had 30 responses. All 30 responses were given an “Overall score” ranked on a 10-point Likert scale, with 0 representing “Totally Unsatisfactory” and “Totally Satisfactory”. In the second group, RAG was done via Approach 2. The second group had 20 responses. Similarly, the 20 responses were given an “Overall score” ranked on the same 10-point Likert scale. In addition, each of the 20 responses was ranked on “Accuracy”, “Conciseness” and “Helpfulness”. The 50 responses were categorized this way because feedback from clinician evaluation of the first group of 30 was used to improve the evaluation process for the second group of 20. The first group of 30 responses were evaluated by all 13 physicians. The second group of 20 responses was evaluated by 8 of the physicians. Evaluators also provided comments on the clinical scenarios and responses. A total of 520 rankings were then analyzed.

### H. Collection and Analysis of Evaluation Reports

Evaluation reports were collected via an online form. Quantitative analysis and associated visualizations were performed in Microsoft Excel Version 16.83. The Real Statistics Resource Pack [16] was used for Interrater Reliability Analysis. For qualitative analysis, evaluators’ comments were compiled as text in a document and coding was performed using Taguette 1.4.1-40-gfea8597 [17]. Thematic analysis and visualization were performed in Python 3.11 [18].

## IV. Results

### A. Quantitative Analysis

Table 1 shows the ranking scores for Response A to C across the 50 responses. These rankings are from the arithmetic mean of each evaluator’s ranking of the 3 responses across the 10 scenarios, rounded up to the nearest whole number for ease of readability. The overall mean ranking across the three models was 7.1 with a standard deviation of 1.4.

**TABLE I.** Mean Response Ranking: Models + Prompt Engineering.

| *Model Response* | *Mean Ranking Score (s.d.)<sup>a</sup>* |
| --- | --- |
| Response A | 6.9 (1.2) |
| Response B | 7.4 (1.3) |
| Response C | 6.8 (1.5) |

<sup>a</sup>. Standard deviation

Table 2 shows the differences in mean ranking scores with a change in RAG approach. The second RAG approach employing chunking via semantic splits, caused an increase in the mean ranking scores of the responses generated by the GPT4-Turbo Preview and Claude Sonnet models as shown in the table.

**TABLE II.** Mean Response Ranking: Models + Prompt Engineering + RAG.

| *Model Response* | *Mean Ranking Score: RAG Approach 1 (s.d.)* | *Mean Ranking Score: RAG Approach 2 (s.d.)* |
| --- | --- | --- |
| Response D | 6.0 (0.6) | 7.2 (1.3) |
| Response E | 6.3 (1.2) | 7.4 (1.1) |

<sup>a</sup>. Standard deviation

Table 3 shows the ranking of the 20 responses in terms of “Accuracy”, “Conciseness”, “Helpfulness” and “Overall Score”. Gemini 1.5 Pro + Prompt Engineering (Response B) elicited the highest rating scores in “Accuracy, “Safety”, “Helpfulness” and “Overall Score”. Claude Sonnet + Prompt Engineering (Response C) and GPT4-Turbo Preview via Open AI Chat Completions API + Prompt Engineering + RAG (Response D) had the same mean rankings, and the highest, for “Conciseness”. GPT4-Turbo Preview via Open AI Assistant API + Prompt Engineering (Response A) had the lowest mean scores for “Conciseness”, the lowest mean rating across the rubric.

**TABLE III.**
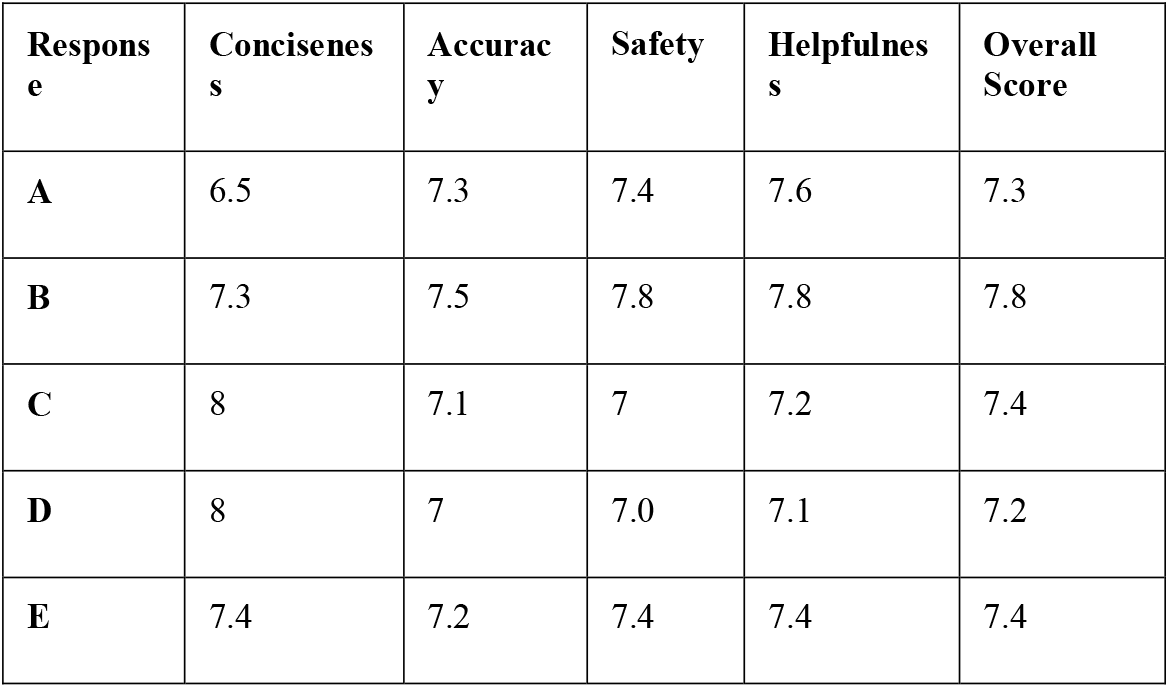
Mean Response Ranking in Accuracy, Conciseness, Safety and Helpfulness.

Gemini 1.5 Pro + Prompt Engineering (Response B) elicited the highest rating scores: 7 or 8 out of 10, at least 85% of the time. GPT 4-Turbo Preview via Open AI Assistant API + Prompt Engineering (Response A) had the second highest ratings: 7 or 8 out of 10, at least 70% of the time. Claude Sonnet + Prompt Engineering + RAG (Response E) had a score of 7 or 8 out of 10, 50% of the time. (Figure 1).

**Fig. 1.**
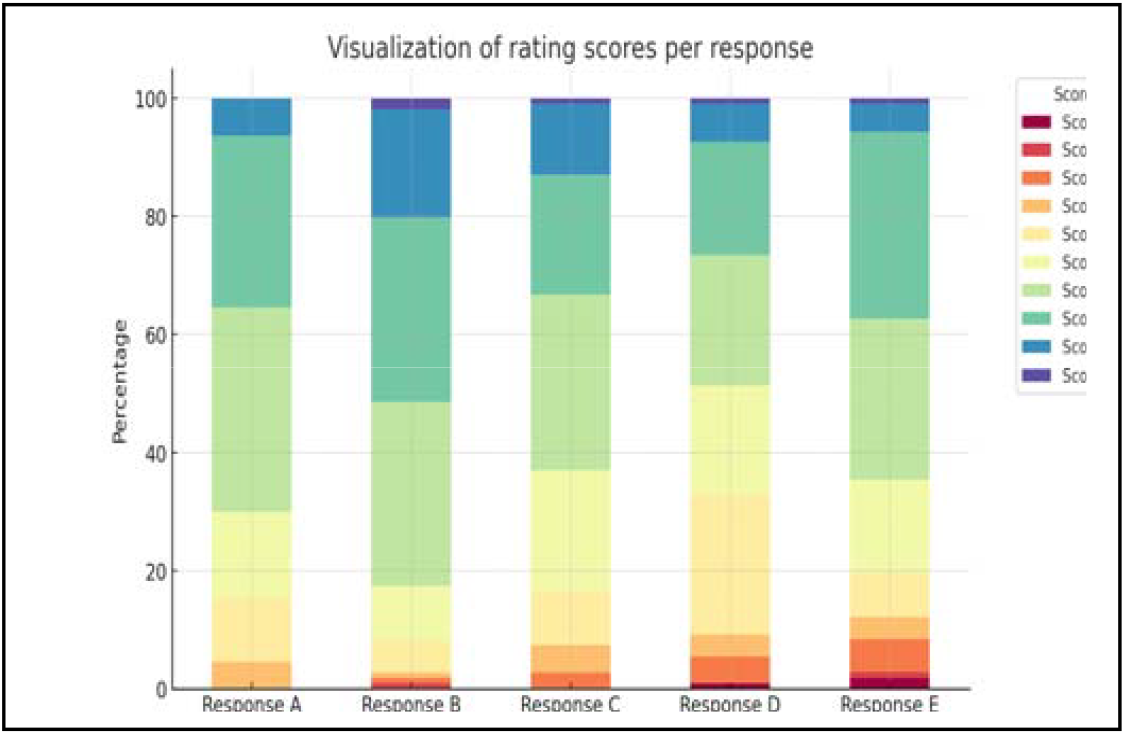
Distribution of rating scores per Response category.

**Fig. 2.**
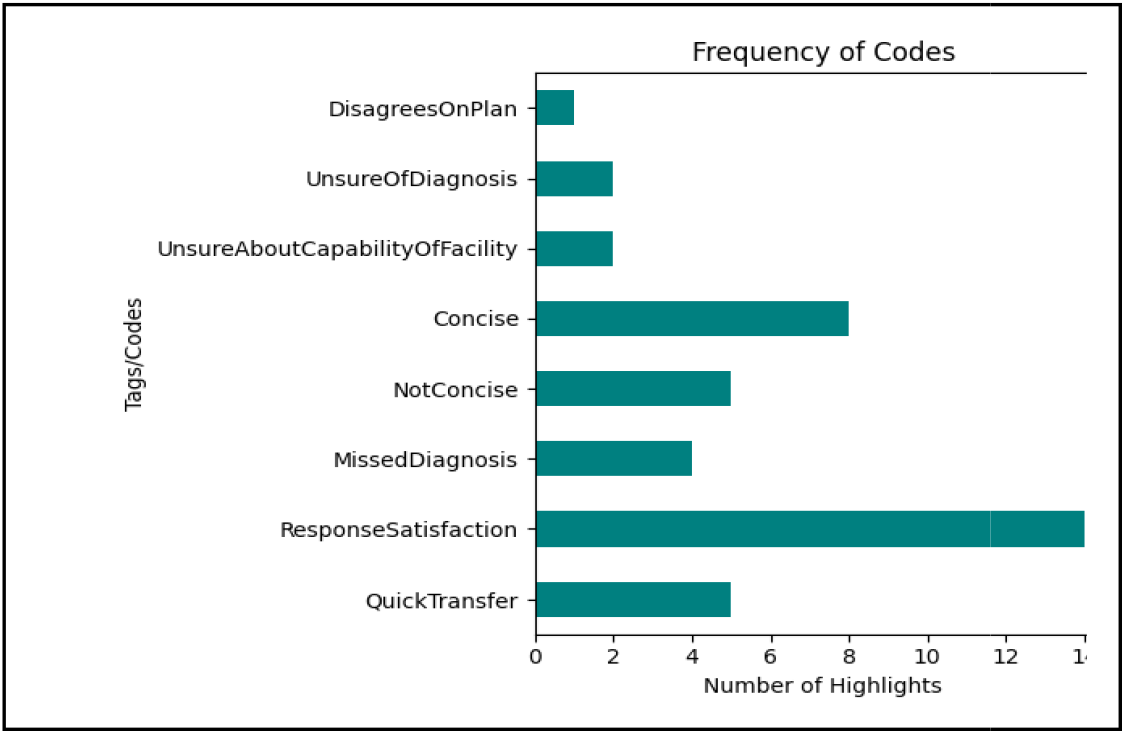
Frequency of Codes in Analysis of Evaluators’ Comments.

Gwet’s AC_2_ score using ordinal weights and a significance level (alpha) of 0.05 was calculated as a measure of interrater reliability. There was a high level of agreement between evaluators, reflected by a Gwet’s AC_2_ score of 0.89.

### B. Qualitative Analysis

8 codes were generated representing recurring viewpoints expressed. Table 3 shows the 8 codes and their descriptions.

***ResponseSatisfaction*** was the most frequently occurring code, indicating numerous instances where the responses were considered satisfactory.

***Concise*** and ***QuickTransfer*** also had significant occurrences, suggesting that the importance of conciseness in responses and the importance of quick transfers were often emphasized. ***MissedDiagnosis*** and ***NotConcise*** were less frequent but notable, indicating areas where responses may have missed critical diagnoses or were not concise enough. outlines the distribution of the codes.

The most commonly occurring codes were grouped into the following themes showing what clinicians considered most in evaluating scenarios and accompanying responses, arranged in descending order of frequency:

- Theme 1. Clarity and Efficiency of Communication: Includes ResponseSatisfaction, Concise, and NotConcise.
- Theme 2. Diagnostic and Management Accuracy: Includes MissedDiagnosis, UnsureOfDiagnosis and DisagreesOnPlan.
- Theme 3. Urgency and Efficiency in Patient Transfer: Includes QuickTransfer and UnsureAboutCapabilityOfFacility.

Table 5 details some of the clinicians’ comments under each of these themes.

**TABLE IV.** Description of Codes.

| Code | Description |
| --- | --- |
| QuickTransfer | Emphasis on transferring or referring the casualty quickly to a health facility |
| ResponseSatisfaction | Expresses positive sentiments about the response |
| MissedDiagnosis | Mentions a diagnosis the response did not provide |
| NotConcise | Expresses dissatisfaction that response is not concise. |
| Concise | Expresses satisfaction that the response is concise |
| UnsureAboutCapabilityOfFacility | Expresses uncertainty and lack of confidence in nearby facilities |
| UnsureOfDiagnosis | Expresses low confidence in the outputted diagnosis |
| DisagreesOnPlan | Disagrees or is unsure of the first aid plan suggested. |

**TABLE V.**
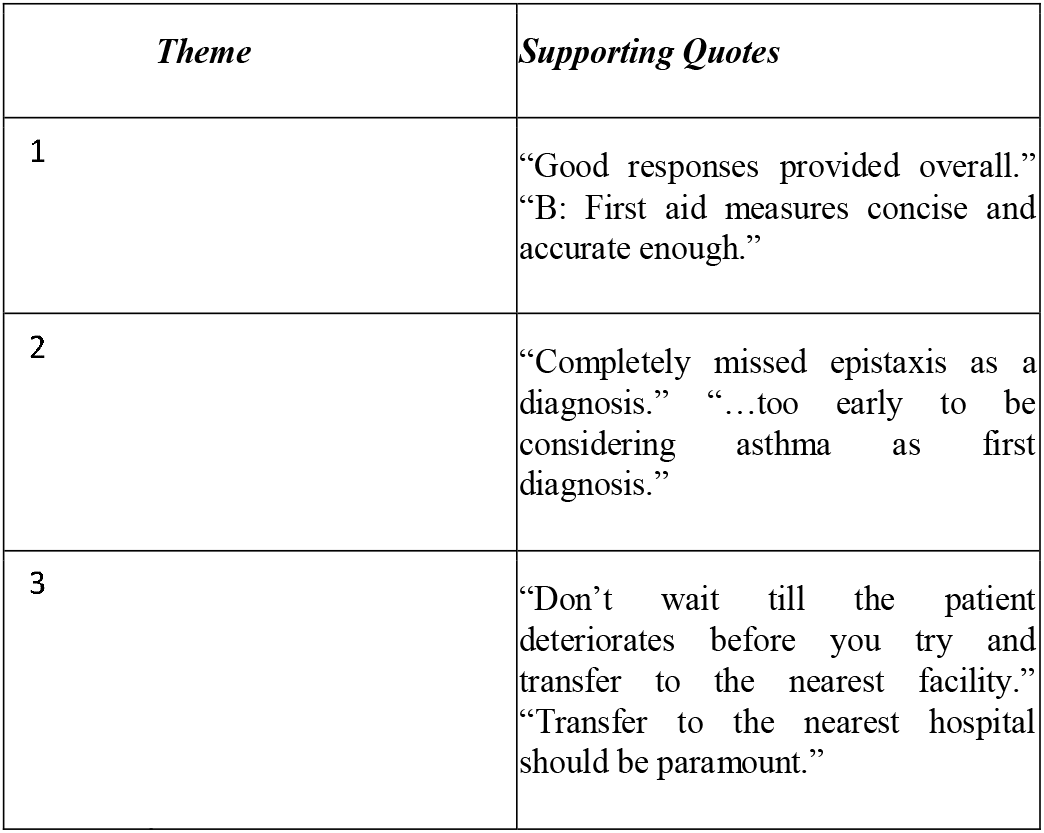
Examples of Evaluators’ comments under the various themes.

### C. Machine Evaluation vs Human Evaluation

BLEU, ROUGE L, ROUGE LSUM and Semantic Similarity scores were computed for the LLM outputs with answers provided from the updated 2021 version of the first aid reference book as the reference text [14]. The semantic similarity score was calculated using cosine similarity with the “all-mpnet-base-v2” transformer model used to generate the vector embeddings for both the LLM output and the reference text. As shown in Figure 3, the scores from the machine evaluation had no correlation with the human ranking scores. Furthermore, LLM outputs with the closest semantic similarity to the reference text did not necessarily have higher human ranking scores.

**Fig. 3.**
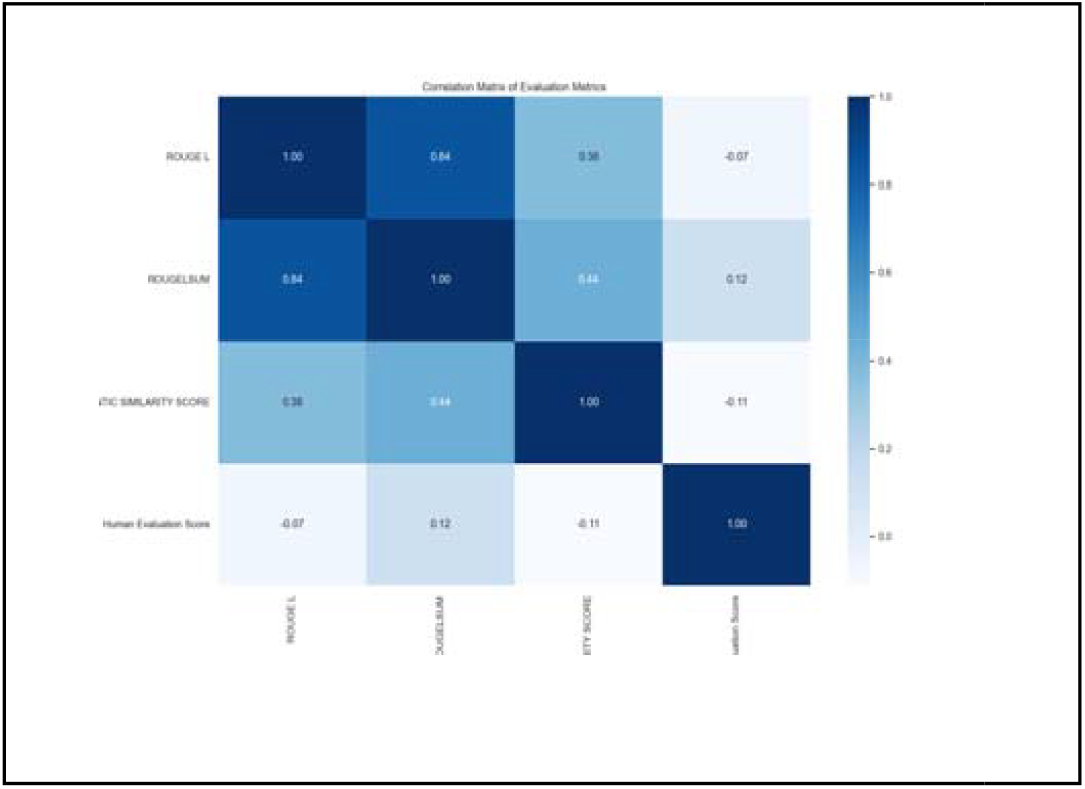
Correlation between auomated metrics and huam evaluation

## V. DISCUSSION

Evaluators were generally satisfied with the diagnosis and first aid instructions outputted by the best performing generalized LLMs combined with moderate prompt engineering as indicated by both the quantitative and qualitative analysis results. This performance by the LLMs is notable considering that they had not had any prior pretraining or finetuning geared for the tasks. Also, the prompting strategy implemented was amongst the simplest with only one-shot inference. Past studies have shown that more sophisticated prompting strategies on generalized LLMs can lead to performances that out-perform state-of-the art, medical LLMs [19]. The best performing model in our study, achieved a mean ranking score of 7/10 which is encouraging. This is a positive finding for resource-constrained settings where the ability to create more specialized, domain-specific models and/or to run them is greatly limited. If generalized models which are often more accessible to wider groups of people, can be made to perform at par/or better than specialized medical LLMs using simpler techniques, then developers in resource-constrained settings can take advantage to develop effective yet cost-efficient applications. An example of such applications is the SnooCODE Red app being developed in Ghana [20]. Figure 4 shows a version of the app in development.

**Fig. 4.**
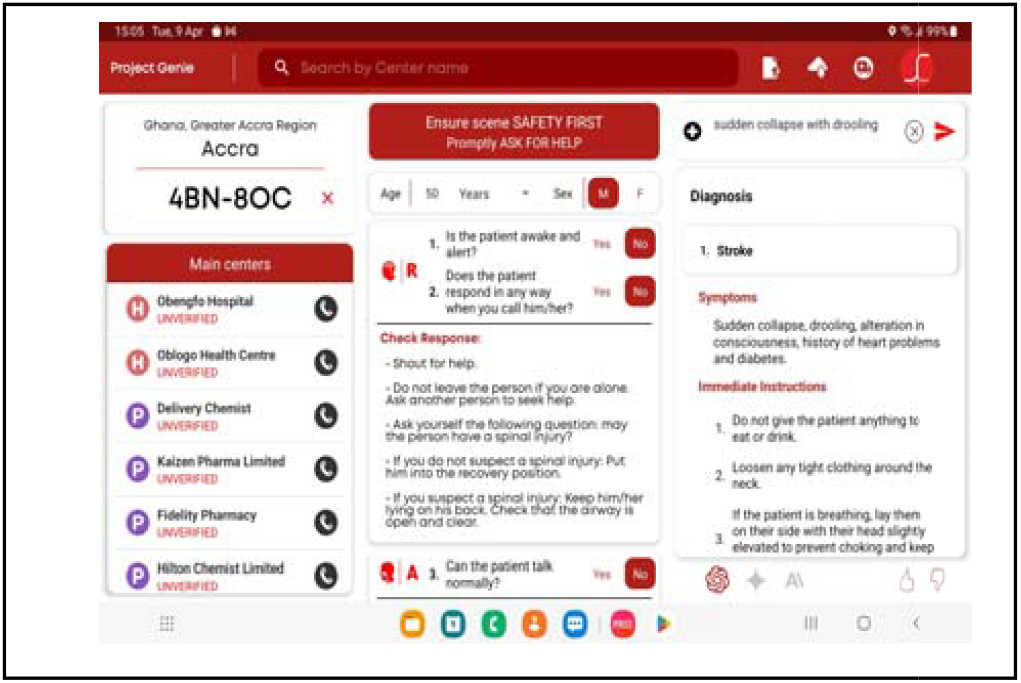
Screenshot of the SnooCODE RED app under development

Though many studies have demonstrated the benefits of RAG in boosting the performance of generalized LLMs in domain specific tasks [12][13][21][22], more emphasis must be placed on RAG technique. Our findings showed that no RAG is better than RAG not done properly. RAG does improve model performance but only with the correct RAG approaches and techniques.

The study also sheds more light on the importance of considering context in the evaluation of LLM performance. This is an area that human evaluators might beat machine evaluators. Our study shows that automatic evaluation metrics did not match human evaluation. Clinician evaluators were not satisfied with responses that did not demonstrate a higher sense of urgency in the transfer of casualties to nearby health facilities even though in the prompt instruction, all the models were informed that EMS was on the way. Responses that instructed that the patient be transported to the nearest health facility even as first aid steps were being instituted were rated as more satisfactory. In contexts with better access to resources, evaluators might not have expressed such a strong concern about waiting for EMS. In many of the rural settings provided in the scenarios with meagre resources, this expression of concern was warranted. This underscores the huge importance of considering contexts in developing LLM-based clinical decision support tools. It is not enough that LLMs pass general medical benchmarks, their performance in different contexts must be evaluated, otherwise responses considered helpful in some settings may not only be unhelpful in other settings, but also harmful.

There are obvious limitations in this study. Firstly, a larger cohort of responses could have been evaluated. Also, a more comprehensive evaluation framework could have been employed. We hope that the feedback obtained can be used to improve LLM outputs for the provided scenarios. We also hope that the insights derived can provide some direction in implementing more detailed and extensive studies of LLM outputs in resource-constrained settings.

## VI. Conclusion

LLM-based first aid assistants have the potential to provide clinically useful instructions in medical emergencies. This is especially helpful in resource-constrained settings where timely access to well-equipped health facilities is often difficult. This potential should be explored further to build applications which may prove life-saving in real-world settings

## Data Availability

All data produced in the present study are available upon reasonable request to the authors.

https://bit.ly/snoocodered-context-matters

